# Modulating amygdala activation to traumatic memories with a single ketamine infusion

**DOI:** 10.1101/2021.07.07.21260166

**Authors:** Or Duek, Yutong Li, Ben Kelmendi, Shelley Amen, Charles Gordon, Madison Milne, John H. Krystal, Ifat Levy, Ilan Harpaz-Rotem

## Abstract

NMDA receptor antagonists have a vital role in extinction, learning, and reconsolidation processes. During the reconsolidation window, memories are activated into a labile state and can be stored in an altered form. This concept might have significant clinical implications in treating PTSD. Using amygdala activity as a major biomarker of fear response, we tested the potential of a single subanesthetic intravenous infusion of ketamine (NMDA receptor antagonist) to enhance post-retrieval extinction of PTSD trauma memories. Post-extinction, ketamine recipients (vs midazolam) showed a lower amygdala and hippocampus reactivation to trauma memories. Post-retrieval ketamine administration was also associated with decreased connectivity between the amygdala and hippocampus, with no change in amygdala-vmPFC connectivity, which suggests that ketamine may enhance post-retrieval extinction of PTSD trauma memory in humans. These findings demonstrate the capacity to rewrite human traumatic memories and to modulate the fear response for at least 30 days post-extinction.

## Introduction

Upon reactivation, a stored memory enters a labile state, in which its content or meaning may be altered. In a process known as reconsolidation, the memory is then stored again in this altered form^1^. This process is especially relevant to post-traumatic stress disorder (PTSD). One of the signature symptom clusters in PTSD comprises overgeneralization of fear and occasional re-experiencing of the traumatic memory with its original emotional intensity and vividness. Memory retrieval, either spontaneous or via deliberate exposure, may open a time window in which adaptive or maladaptive memory modifications can promote either attenuation or persistence of PTSD symptoms ^1,2^.

In the laboratory, fear learning is typically modeled by simple paradigms, in which neutral cues are paired with aversive outcomes (e.g electric shocks) and acquire aversive value^3^. Following fear acquisition, repeated presentation of cues in the absence of outcome leads to the extinction of fear responses. But these responses usually return, either spontaneously or following further exposure to the cue or the aversive outcome ^4,5^, suggesting that the original memory is not substantially altered. Extinction is likely achieved through inhibition of amygdala fear responses by the ventromedial prefrontal cortex (vmPFC)^6^. However, if extinction training is conducted within the reconsolidation window, attenuation of fear can be long-lasting ^7,8^ and likely involves modification of the original memory ^9^. Such post-retrieval extinction does not appear to rely on prefrontal mechanisms ^10,11^, and may directly target the memory trace in the amygdala ^1,12,13^. Functional connectivity between the amygdala and hippocampus may also play an important role in the processing of aversive memories ^11,14,15^.

Extinction may be further enhanced with the use of ketamine, a non-competitive N-methyl-D-aspartate glutamate receptor (NMDAR) antagonist. The NMDAR has a key role in learning, extinction, and reconsolidation in both animals and humans ^16–18^. Moreover, accumulating evidence suggests that ketamine, in sub-anesthetic doses, promotes neurogenesis ^19,20^, cell proliferation^21^, and synaptogenesis ^20,22^, all of which are important in reconsolidation processes ^23,24^.

While post-retrieval extinction holds considerable clinical promise for the treatment of PTSD, it is not yet known whether its effect translates from laboratory-based fear learning to real traumatic memories^25^, and from healthy participants to those suffering from trauma-related psychopathology ^26,27^. Here we explored the potential of a single subanesthetic intravenous infusion of ketamine (0.5mg/kg over 40 min) to enhance post-retrieval extinction of real traumatic memories. To control for the subjective effects of ketamine, we randomized individuals with PTSD to receive an infusion of either ketamine or the benzodiazepine midazolam. As ketamine was found to promote reconsolidation processes, we combined the infusion with a reconsolidation-focused four-day exposure-based therapy. Our aim was to identify neural biomarkers for reconsolidation of the original traumatic memory for a potential transitional clinical trial. Our main hypothesis focused on the amygdala; we predicted greater post-retrieval reduction in amygdala reactivity to trauma cues in individuals with PTSD who received ketamine, compared to those who were administered midazolam. As the target engagement mechanism was of reconsolidation and not extinction, we expected no change in vmPFC activation or its functional coupling with the amygdala. Lastly, we hypothesized that if ketamine produces greater post-retrieval extinction than midazolam, this effect will also be associated with a reduction in functional connectivity between the amygdala and the hippocampus.

## Method

### Participants

Of the 125 participants screened, 30 were eligible and 28 were randomized (two did not randomize due to lost contact). One subject voluntarily left the experiment immediately after the infusion and another was excluded from the analysis due to diagnostic uncertainty, resulting in 26 participants who were included in the analyses (mean age =39.7, SD = 10.8, range=26-66; 13 subjects per group; females (n=10), males (n=16)). See Attached CONSORT figure.

Exclusion criteria included a diagnostic history of bipolar disorder, borderline personality disorder, obsessive-compulsive disorder, schizophrenia or schizoaffective disorder, or current psychotic features as determined by the Structured Clinical Interview for DSM-IV (SCID)^28^; dementia was also an exclusion criterion, as were current suicide risk, moderate or higher severity of substance use disorder in the 3 months prior to randomization, and history of mild-to-severe traumatic brain injury (TBI). Participants who were currently engaged in trauma focus therapy were also ineligible to participate in the study.

PTSD diagnosis was established using the Clinician-Administered PTSD Scale (CAPS-5)^29^. The PTSD Checklist for DSM-5 (PCL-5) was used to monitor change in PTSD symptoms over time^30^. Patients were excluded for acute medical illness based on medical history, physical examination, and screening laboratory test values. We screened for possible cardiac issues using EKG.

### Procedure

Eligible participants completed an imagery-development procedure in which they were asked to describe the traumatic event associated with their PTSD (Criteria A), as well as a sad event and an event in which they felt relaxed (referred to as neutral). The imagery scripting procedure followed a procedure presented by Sinha et al.^31^. Subjects were asked to describe the events in as much detail as possible. They were then asked to circle at least three physiological responses corresponding to each specific event. Using this information, we developed a 120-second audiotape script of each event, narrated by a male member of the research staff.

24 hours post infusion, participants began daily exposure-based therapy that included imaginal exposure (i.e. reviving the traumatic event in the patient’s imagination) and in-vivo exposure (i.e. confronting unpleasant stimuli that the patient started to avoid after the traumatic event). The entire study procedure (including imaging sessions) lasted 7 days. On day 1, participants underwent a session that included psychoeducation, life history, and generation of a scale to gradually confront harder and harder situations during the in-vivo exposure practice, based on prolonged exposure psychotherapy^32^ procedure. On day 2, a baseline MRI scan was conducted which included T1 MPRAGE, 10-minute resting state, DTI, and listening to the 3 scripts (traumatic, sad, and neutral, each 120 seconds long). Each script was repeated three times for a total scan length of 18 minutes. Following script replay (scripts were replayed to each subject in a constant order), while the trauma memory was in a labile state, infusion of either ketamine (0.5mg/kg) or midazolam (0.045mg/kg) began inside the MRI for 40 minutes.

From day 3 to day 6, participants underwent trauma memory exposure sessions in which they activated their original traumatic memory using free recall as well as in-vivo exposure, based on the participants’ exposure hierarchy. This procedure was done with a clinical psychologist and focused on reviving the memory and reconsolidation of the memory in a more processed and organized state.

On day 7, participants were scanned again, using a similar procedure to day 2 but without the infusion. A self-report questionnaire assessing participants’ current PTSD symptomatology was also taken. Lastly, 30 days post-treatment, a follow-up session was conducted, assessing both neural responses to the same scripts and PCL-5 scores. For an illustration of the procedure, see figure 1.

**Figure 1:**
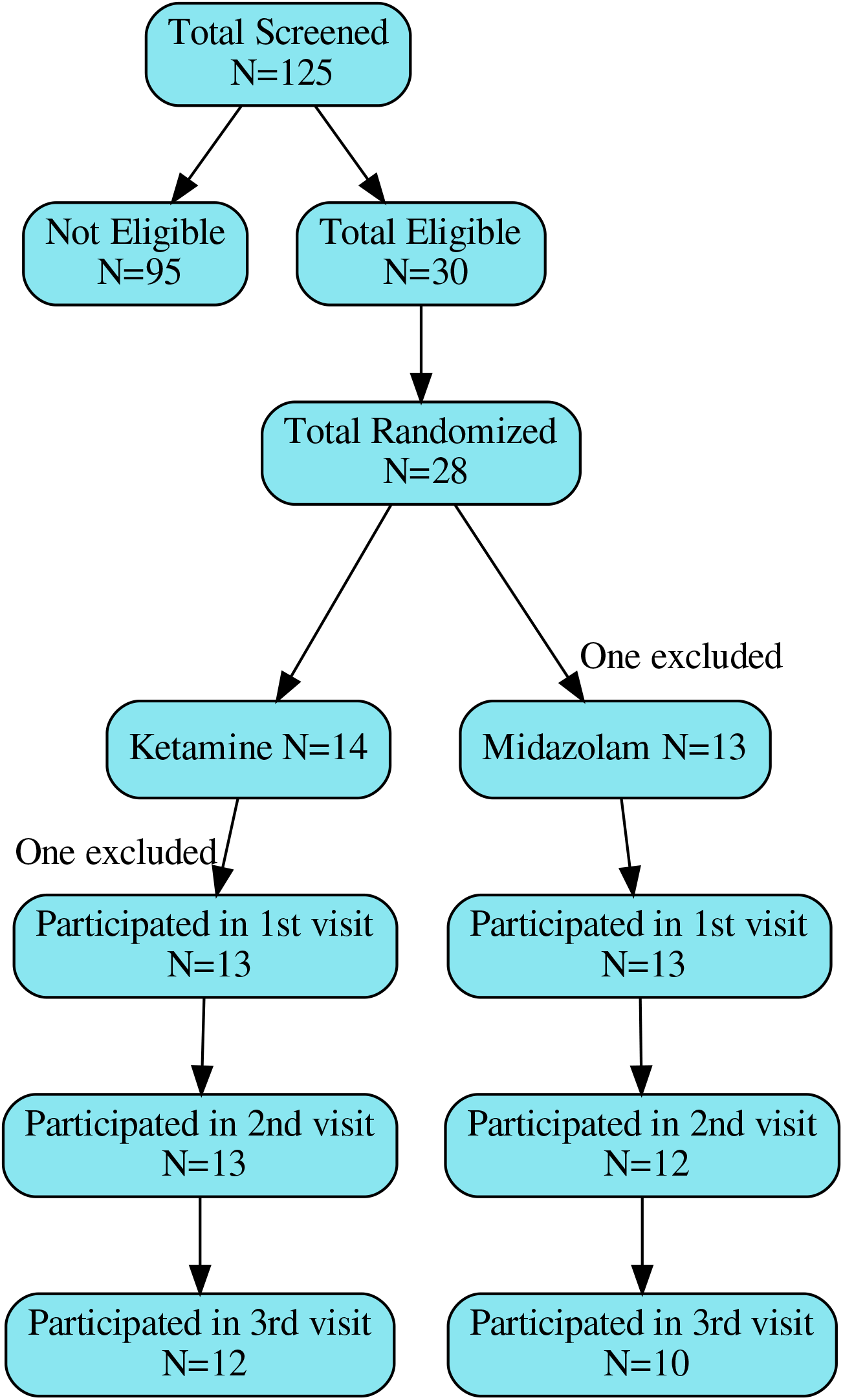
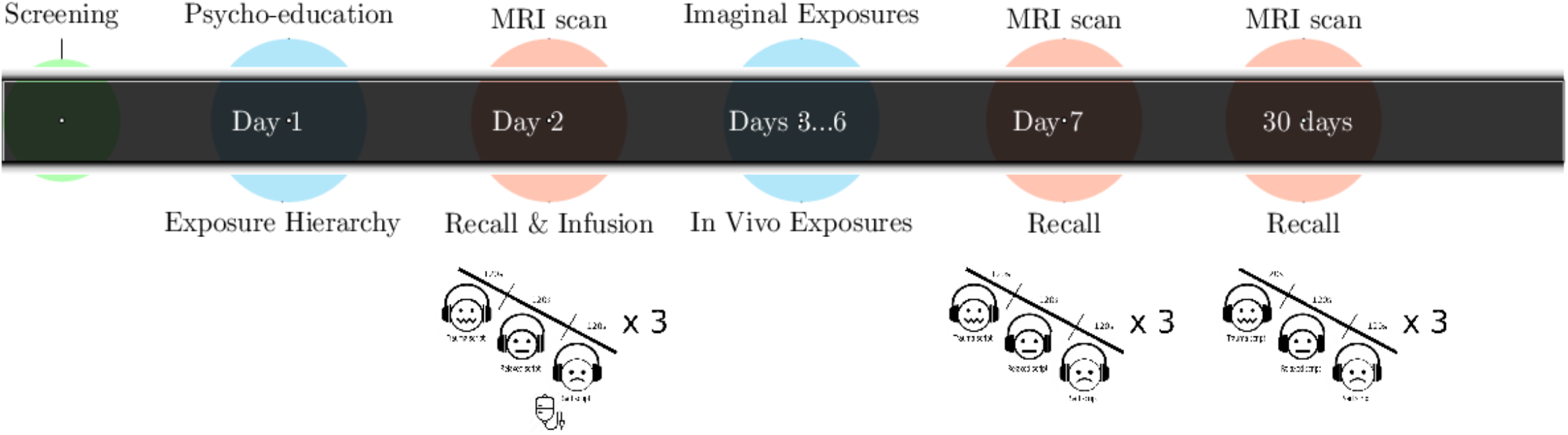
Illustration of the procedure. Days 3..6 = days 3,4,5 and 6 in which the PE sessions were conducted. Recall procedure inside the magnet included 3 rounds of each audio recall script (traumatic, sad, and neutral, 120 seconds each)

### Skin Conductance Response

Skin conductance data were collected with shielded Ag–AgCl electrodes, filled with standard NaCl electrolyte gel, attached to the middle phalanges of the second and third fingers of the left hand. The electrode cables were grounded through a radio frequency filter panel. The skin conductance signal was amplified and recorded with a BIOPAC Systems skin conductance module connected to a computer.

Skin conductance response (SCR) was analyzed using neuroKit2^37^. We used a method that averages SCR peaks in a specific time slot, rather than searching for one peak as in the event-related analysis (when an event is shorter than 10 seconds). This method was more suitable for longer trials, such as listening to an audio clip ^37^.

### MRI Scans

MRI data were collected with a Siemens 3T Prisma scanner, using a 32-channel receiver array head coil. High-resolution structural images were acquired by Magnetization-Prepared Rapid Gradient-Echo (MPRAGE) imaging (TR = 1.9 s, TE = 2.77 ms, TI = 900 ms, flip angle = 9°, 176 sagittal slices, voxel size = 1 ×1 × 1 mm, 256 × 256 matrix in a 256 mm FOV). Functional MRI scans were acquired while the participants were listening to the narrated scripts, using a multi-band Echo-planar Imaging (EPI) sequence (multi-band factor=4, TR= 1000 ms, TE= 30ms, flip angle=60°, voxel size = 2 × 2× 2 mm, 60 2 mm-thick slices, in-plane resolution = 2 × 2 mm, FOV= 220mm).

### Neuroimaging analyses

#### Preprocessing

Data were preprocessed with Fmriprep, version 1.5.6^38^. For a complete preprocessing procedure please see supplement 3. All images were motion-corrected and slice-time corrected, aligned to T1, and then to MNI space. Analysis of the functional MRI data included the following regressors: 6 movement variables (translation and rotation), framewise displacement, deviation of successive difference images (DVARS), the first 6 anatomical components based noise correction (CompCor;^39^). Analysis of activation included a 6mm full width at half minimum (FWHM) smoothing of the images (smoothing was not included in pattern analysis). For a full description of preprocessing, please see supplement 3.

#### Activation level analysis

For each of the three fMRI sessions, a first-level analysis comparing the first 60 seconds of the first traumatic script vs. the first 60 seconds of the first neutral script was conducted using FSL 6.0.3^40^, through a Nipype pipeline^41^. To avoid capturing habituation effects, we chose to focus on the first 60 seconds of the first presentation of each script.

The resulting contrast of parameter estimates (COPE), after z-scoring, were then used to assess specific regions of interest (ROIs) in line with our preliminary hypothesis (i.e., the amygdala, hippocampus, and ventromedial prefrontal cortex (vmPFC)).

Each subject’s COPE was masked using the relevant ROI and averaged across all voxels of that region. The extraction of activation for specific ROIs was conducted using Nilearn^42^. Statistical analyses were conducted using python, and Bayesian comparison of groups was done using the pyMC3 package^36^. Lastly, we compared group differences in the same brain regions during the sad (vs. neutral) scripts. This was done to account for an alternative explanation: that the results we see are general to all negative emotions, and not specific to trauma-related ones.

#### Connectivity analysis

In order to assess connectivity between different ROIs, we used the DiFuMO atlas^43^, including 256 regions, among them the amygdala, hippocampus (anterior and posterior), and ventromedial prefrontal cortex (vmPFC; and vmPFC anterior; see supplement 1 for region maps). We employed the python package Nilearn^42^ to extract time series, using low-pass filtering of 0.1Hz and high-pass filtering of 0.01Hz, and regressing out framewise displacement, CSF, white-matter volume, 6 rotation and translations variables, and first six anatomical components based noise correction (CompCor). Then, we extracted the correlation between those regions from the first 60 seconds of the first traumatic script. We applied pyMC3^36^ to compare changes in correlation between the relevant ROIs (amygdala, anterior and posterior hippocampus, anterior vmPFC, and vmPFC). Lastly, we conducted the same procedure on the first 60 seconds of the sad script (as we did in the activation analysis), in order to test whether these effects are related to negative emotions in general or to trauma-related ones specifically.

#### Statistical approach

In accordance with recent guidelines and findings ^33–35^, analysis in this manuscript was based on a Bayesian approach. All analyses were conducted using pyMC3, a probabilistic programming package in python^36^.

The analysis of PTSD symptom change was based on multilevel modeling, with subjects as a random variable (i.e. different intercept for each subject). Before analysis, PCL-5 scores were scaled for easier interpretation. A normal distribution with mean=0 and sd=1 was set as prior to subject intercept and time variables (to test the different times we set three priors, one for each time point). A normal distribution of mean=0 and sd=0.5 was set for group difference. Lastly, to analyze the effect of time, we subtracted the posterior distribution of each time point from the distribution of time point zero (before treatment) and assessed the probability that this new distribution was less than zero.

Analysis of SCR employed the same concept, using subjects as a random variable. A normal distribution with mean=0 and sd=1 was set to the intercepts, as well as a time variable. A normal distribution with mean=0 and sd=0.5 was set as prior to the group variable.

Analysis of fMRI compared the groups at each time point, assessing the level of difference in activation (or connectivity) between the groups before treatment, after treatment (7-days after infusion), and at 30-day follow-up. Full analysis scripts can be found here: https://github.com/orduek/KPE.

## Results

Twenty-six subjects suffering from PTSD received daily sessions of exposure therapy for four consecutive days, following infusion of either ketamine (n=13) or midazolam (n=13). Subjects participated in three functional MRI (fMRI) sessions: before infusion, at the end of treatment (7 days after the infusion), and at 30-day follow-up. During these sessions, subjects listened to a narrated version of their traumatic memory, as well as sad and neutral ones (see Methods). Skin conductance responses (SCR) to each memory were recorded, and a self-report questionnaire was used to assess PTSD symptom severity in each session.

### Changes in neural activation

Based on our hypotheses, the main neural analysis focused on activation and connectivity patterns of the amygdala, the hippocampus, and the vmPFC. These regions of interest (ROIs) were defined using Neurosynth^44^. In each ROI, we contrasted activation during the traumatic script with activation during the neutral script at each timepoint. Our statistical approach was based on a Bayesian model with non-informed priors. This means that we have created a new probabilistic model for each timepoint, using a model similar to linear regression with group as a variable predicting activation. Thus, the result was the group difference for that specific time point.

### Amygdala

Activation difference for traumatic vs. neutral memories was lower in the ketamine group than in the midazolam group 7 days after infusion, with mean activation of ketamine group -0.15 (sd=0.35;N=13), mean activation of midazolam group 0.18 (sd=0.3; N=12) and mean difference of the posterior distribution: -0.346,sd=0.14, 90%HDI^1^ [-0.586,-0.109]). This difference remained significant at the 30-day follow-up (ketamine: -0.14 (sd=0.35;N=12), midazolam: 0.15 (sd=0.38;N=10), mean difference from posterior distribution: -0.294, sd=0.173, 90%HDI [-0.572, -0.005]). No group difference was found before treatment (ketamine: 0.01 (sd=0.40;N=13), midazolam: 0.09 (sd=0.38; N=13), mean difference from posterior distribution: -0.083, sd=0.16, 90%HDI [-0.356, 0.186]; see figure 2a,b). There was no group difference in amygdala activation during the sad imagery scripts at any timepoint (see supplement 2).

**Figure 2:**
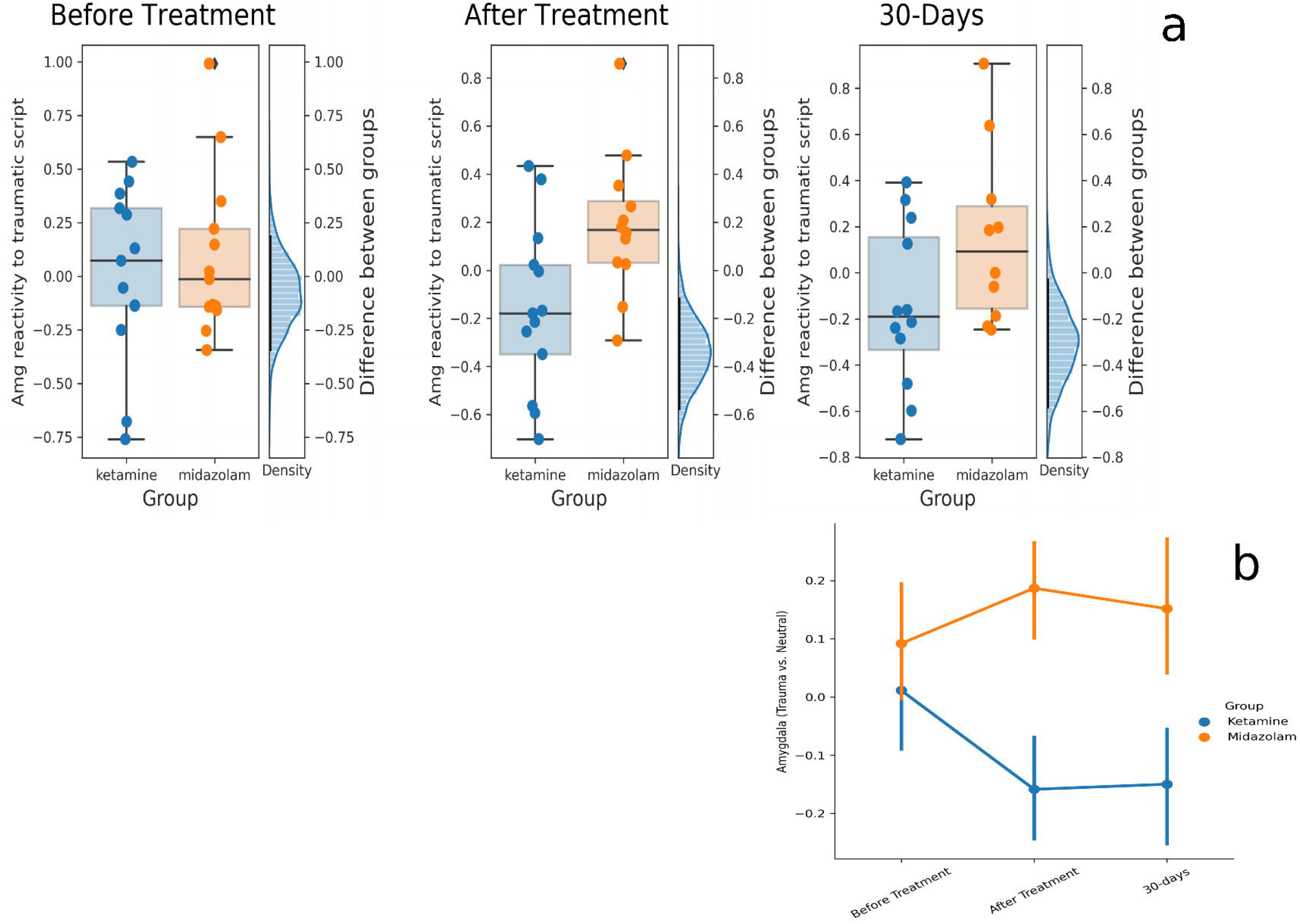
**(a)** Differences between ketamine and midazolam groups in amygdala reactivity to trauma vs. neutral script across the three time points. Each dot is a participant. The distribution on the right of the graph is the posterior distribution of the difference between the groups. The black line is the 90% HDI. **(b)** Average amygdala reactivity to trauma vs. neutral in the ketamine group (orange) and the midazolam group (blue) in the three-time points. Error bars represent SEM.

### Hippocampus

Ketamine group activation in the hippocampus (trauma vs. neutral) was marginally lower in the ketamine (mean=-0.11, sd=0.40; N=13) compared to the midazolam (mean=0.18, sd=0.53;N=12) group, with mean difference from posterior distribution -0.290 (sd=0.199), 90%HDI [-0.620, 0.028]. There was no group difference in the 30-day follow-up (ketamine: -0.15 (sd=0.38; N=12), midazolam: 0.01 (sd=0.42; N=10), mean difference of posterior distribution - 0.177, sd=0.189, 90%HDI [-0.491, 0.122]). At baseline, the level of activation in both groups was also similar (ketamine: 0.02 (sd=0.29; N=13), midazolam: -0.03 (sd=0.26; N=13), mean difference of posterior distribution 0.062, sd=0.12, 90%HDI [-0.14, 0.25]). These results suggest that the effect on the hippocampus was transient, as opposed to the long lasting effect in the amygdala (figure 3). There were no group differences in hippocampus activation of the sad imagery script at any time point (see supplement 2).

**Figure 3:**
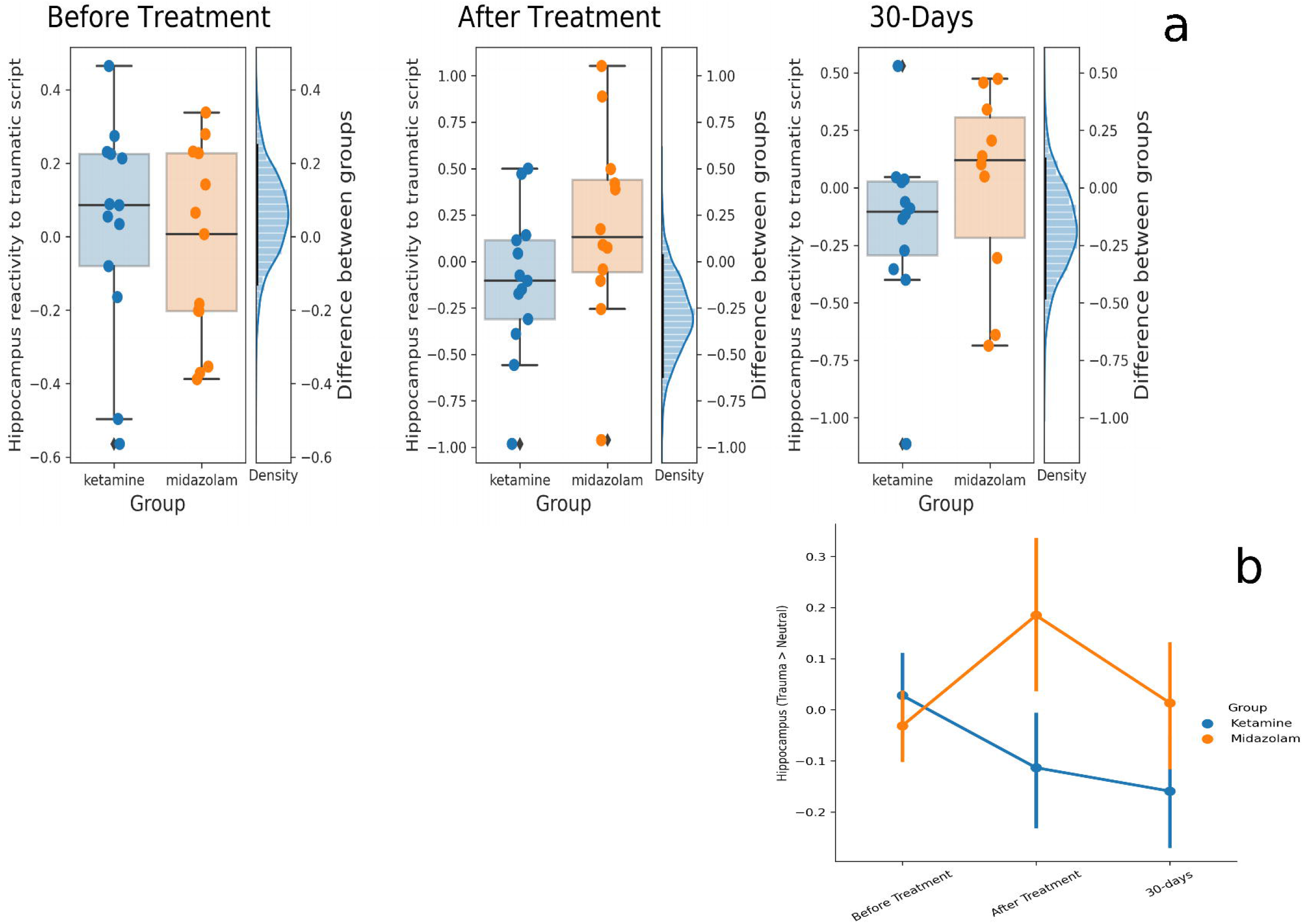
**(a)** Differences between ketamine and midazolam groups in hippocampus reactivity to trauma vs. neutral script across the three time points. Each dot is a participant. The distribution on the right of the graph is the posterior distribution of the difference between the groups. The black line is the 90% HDI. **(b)** Average hippocampus reactivity to trauma vs. neutral in the ketamine group (orange) and the midazolam group (blue) in the three time points. Error bars represents SEM

### vmPFC

We found no group differences in vmPFC activation to traumatic imagery scripts (compared to neutral) at any timepoint. There was no group difference in vmPFC activation during the sad imagery scripts compared to the neutral ones at any of the three timepoints. For a full description of the vmPFC activation results, please see supplement 2.

### Changes in skin conductance response (SCR)

To further validate fear activation to trauma memory, psychophysiological response to traumatic memory was assessed using SCR to the memories. The ketamine group (mean=0.2, sd=0.3, N=11) showed lower SCR response to the traumatic imagery script compared to the midazolam group (mean=1.2, sd=1.6,N=8) 7 days after infusion (mean of posterior distribution=-1.004, sd=0.56, 90%HDI [-1.89,-0.06]; Figure 4). This difference between ketamine (mean=0.15, sd=0.23, N=10) and midazolam (mean=0.54, sd=0.77, N=7) remained marginally significant at the 30-day follow-up (mean of the posterior distribution of the difference: -0.38, sd=0.29, 90%HDI [-0.84, 0.09]). There was no group difference between ketamine (mean=0.37, sd=0.73, N=12) and midazolam (mean=0.62, sd=0.71, N=11) in SCR before the treatment (mean of the posterior distribution of difference: -0.25, sd=0.33, 90%HDI [-0.78, 0.30]).

**Figure 4:**
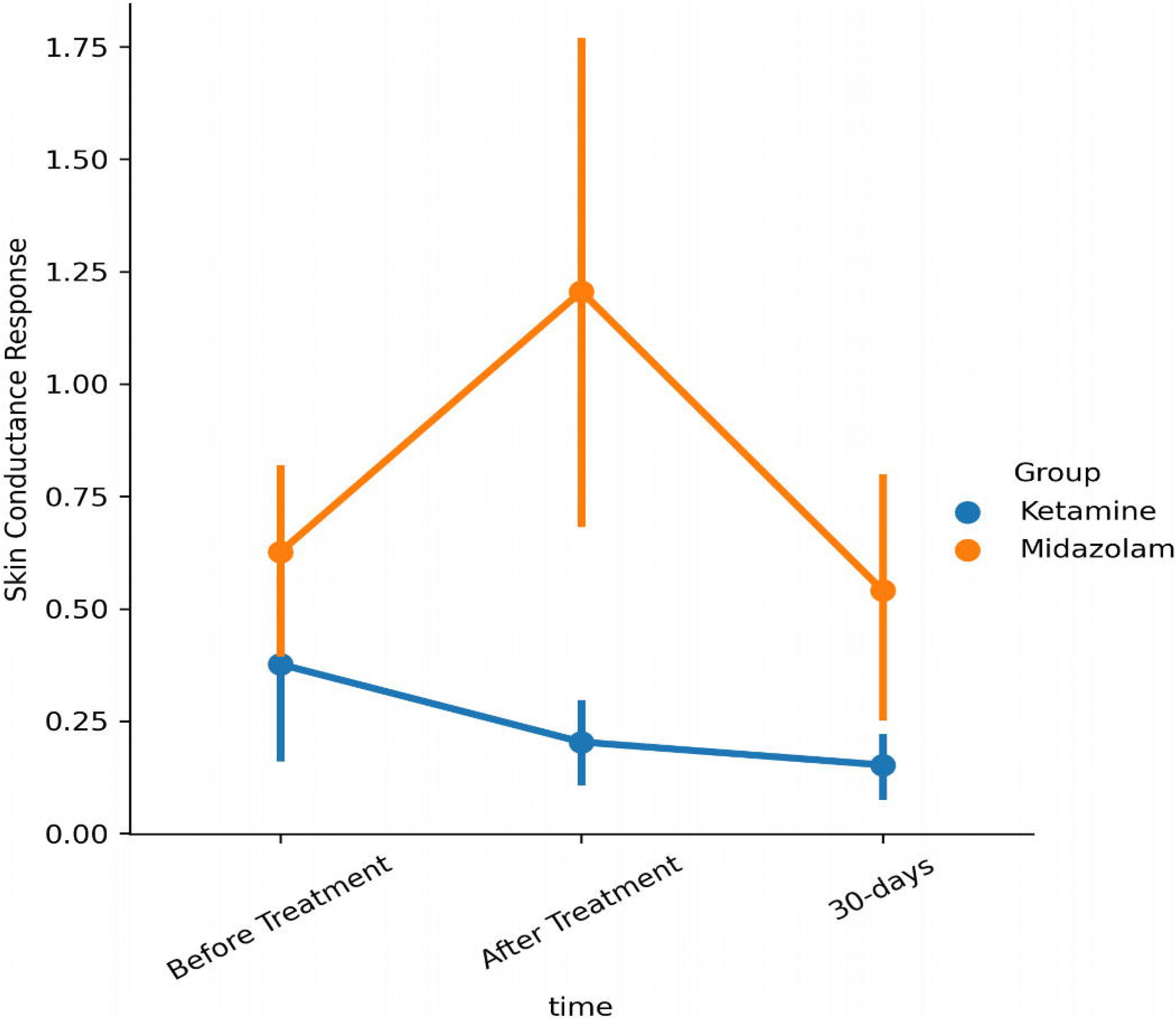
Changes in SCR (Error bars are standard error of the mean)

#### Changes in connectivity

Reduced amygdala activity, in the absence of increased vmPFC activity, is consistent with a direct effect of reconsolidation-based extinction on the amygdala. This differs from what is expected in standard extinction, in which vmPFC (or, more broadly, the prefrontal cortex) is thought to inhibit the amygdala. Thus, while increased connectivity between the vmPFC and the amygdala is generally expected in extinction, in reconsolidation-based extinction there should be no change in connectivity between those regions. Conversely, functional connectivity between the amygdala and hippocampus is expected to change, as reduced connectivity was found to be associated with extinction in the reconsolidation window. Based on these hypotheses, we examined the correlation between amygdala and vmPFC activity, as well as the correlation between amygdala and hippocampus.

### Amygdala-vmPFC

Functional connectivity (FC) between amygdala and vmPFC was not significantly different between the groups 7 days after the infusion, with average FC in the ketamine group: 0.40 (sd=0.21;N=13) and average FC in midazolam group of 0.31 (sd=0.22;N=12); mean difference of posterior distribution was 0.084, sd=0.096, 90%HDI [-0.078,0.233]. The ketamine group showed marginally lower FC at the 30-day follow-up (mean FC in the ketamine group: 0.34, sd=0.25; N=12, mean FC in midazolam group: 0.48, sd=0.15; N=10). Mean difference of posterior distribution: -0.14, sd=0.1, 90%HDI [-0.306, 0.025]). The groups’ FC did not differ before treatment (ketamine: 0.55 (sd=0.22; N=13), midazolam: 0.47 (sd=0.23; N=13), mean difference: 0.069, sd=0.092, 90%HDI [-0.075, 0.222]). Figure 5 depicts the average connectivity of each group at all time points. Moreover, analysis revealed a time effect with lower amygdala-vmPFC connectivity after treatment. To assess the statistical difference, we compared the posterior distribution of each timepoint (after treatment, and 30-day follow-up) to the posterior distribution before treatment. The overlap between the posterior distribution before and after treatment was 0.006, which suggests that the observed decline is significant. Using the same method on the 30-day follow-up revealed an overlap of 0.04, which also shows a decline in the 30-day follow-up connectivity between amygdala and vmPFC compared to before treatment. Post hoc analyses of the time effect showed that in both groups there was a moderate effect size in declined amg-vmPFC FC 7 days after infusion (midazolam: t(22)=1.7, cohen’s-d=0.6, p=0.1; ketamine: t(24)=1.7, cohen’s-d = 0.66, p=0.1). However, at 30-day follow-up the ketamine group showed a further steep decline in amg-vmPFC FC (t(22)=2.15, cohen’s-d=0.86, p=0.04), while in the midazolam group connectivity returned to pre-treatment level (t(20)=-0.11, cohen’s-d=0.04, p=0.9). Comparing amygdala-vmPFC functional connectivity during the sad imagery script revealed no differences between the groups at all three timepoints (see supplement 2).

**Figure 5:**
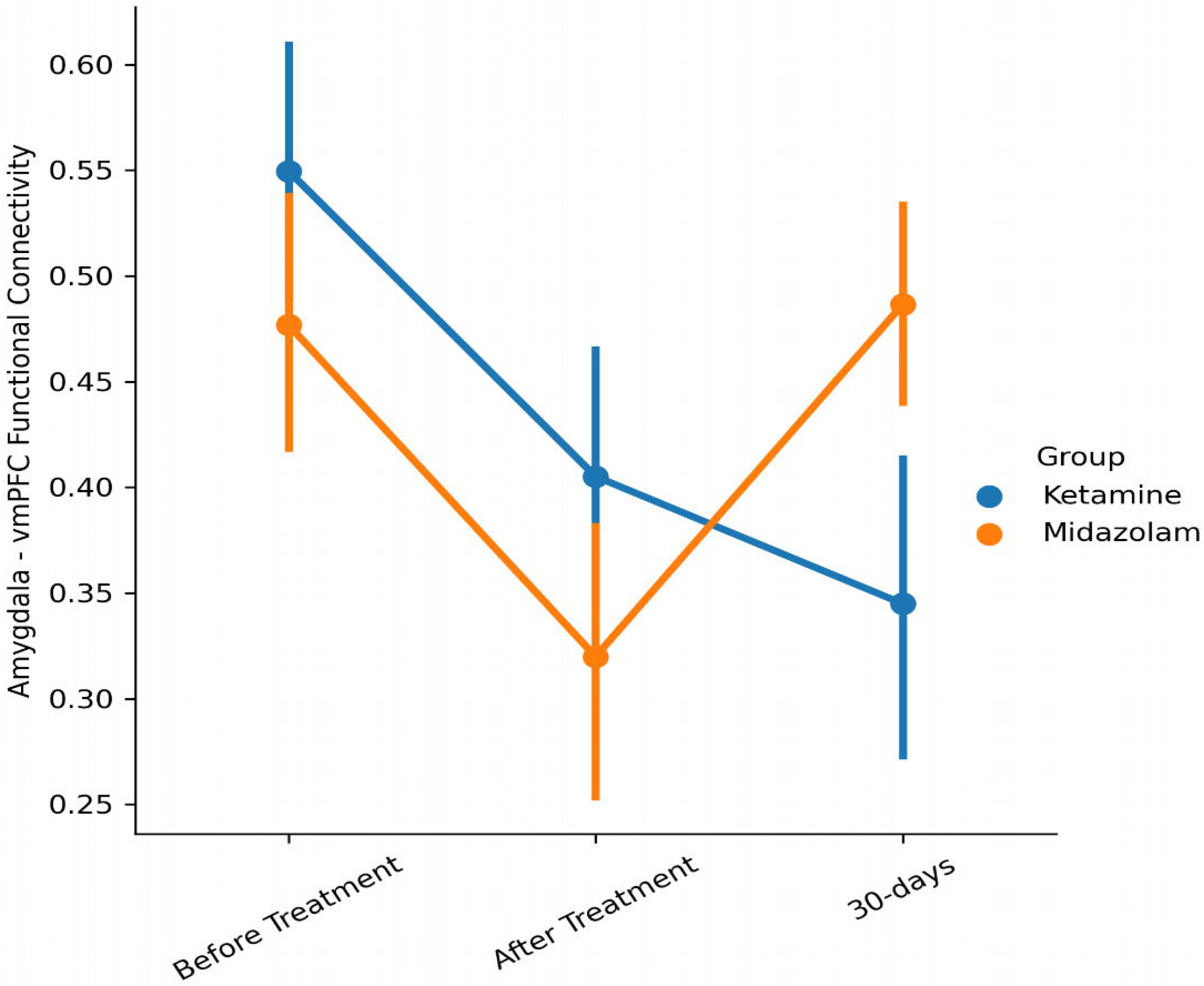
Average connectivity between the amygdala and vmPFC in the ketamine and midazolam groups across the different time points. Error bars represent the standard error of the mean.

### Amygdala - Hippocampus

Functional connectivity (FC) between amygdala and posterior hippocampus 7 days after infusion was significantly lower in the ketamine group (mean 0.16, sd=0.32; N=13) than in the midazolam group (mean 0.43, sd=0.18; N=12), with mean difference of posterior distribution: - 0.274, sd=.11, 90%HDI [-0.46, -0.09]. At 30-days follow-up we found no difference between the ketamine (mean FC: 0.37, sd=0.26; N=12) and midazolam (mean FC: 0.29, sd=0.35; N=10) groups (mean difference of posterior distribution: 0.08, sd=0.15, 90%HDI [-0.17, 0.31]. At baseline, both ketamine (mean FC: 0.31, sd=0.23; N=13) and midazolam (mean FC: 0.23, sd=0.15; N=13) groups had similar FC (mean difference of posterior distribution: 0.007, sd=0.08, 90%HDI [-0.06, 0.21]). Figure 6 shows the average functional connectivity between the amygdala and posterior hippocampus. As with hippocampus reactivity, these results suggest a transient effect. Lastly, using a multilevel model revealed no effect for time.

**Figure 6:**
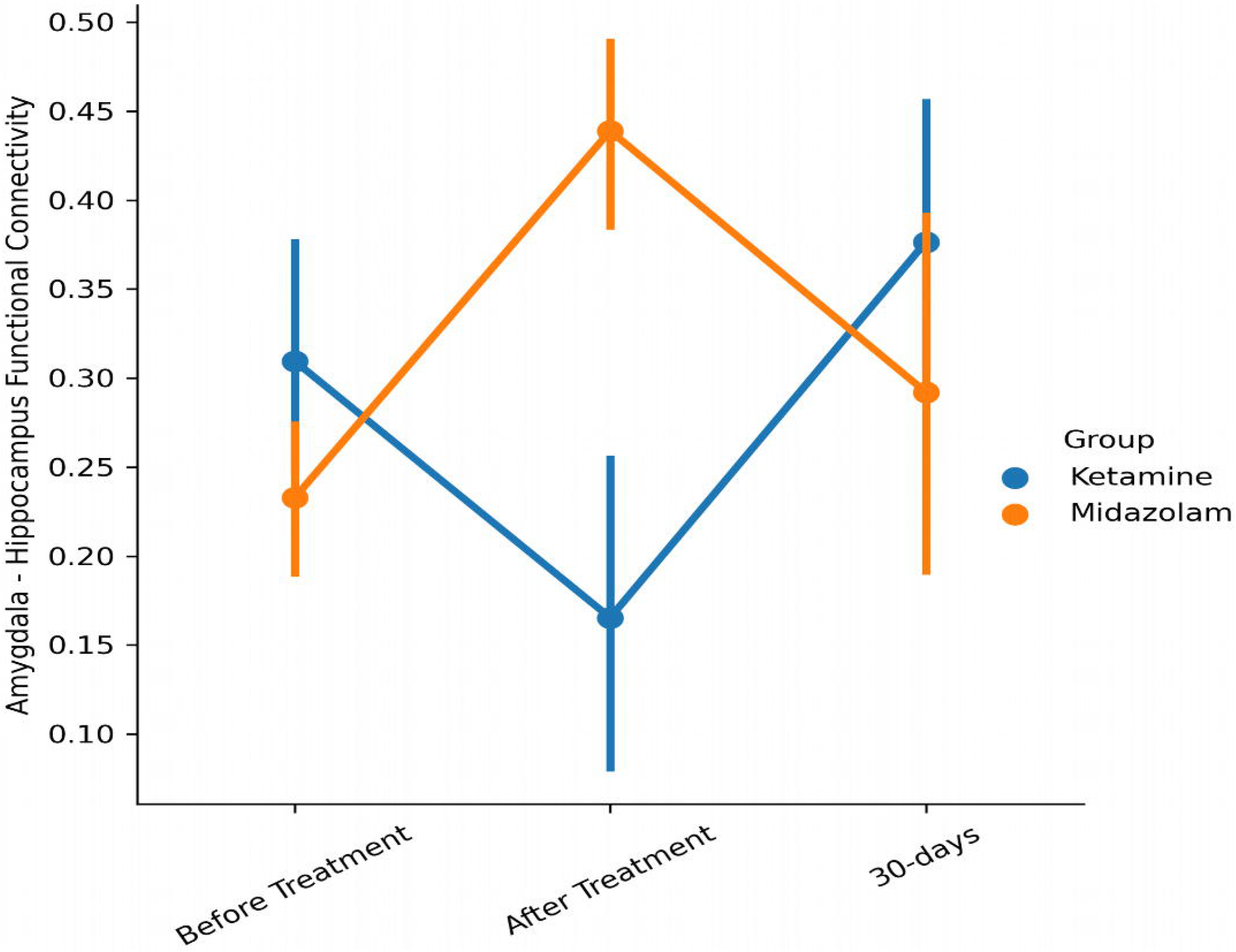
Average connectivity between the amygdala and posterior hippocampus in the ketamine and midazolam groups across the different time points. Error bars represent the standard error of the mean.

Comparing amygdala-posterior hippocampus functional connectivity during the sad imagery script revealed no differences between the groups in all three time points (see supplement 2).

### Changes in PTSD symptoms

Although the main purpose of this investigation was to establish a reliable biomarker for the development of a potential clinical intervention, we also tracked changes in PTSD symptoms during the study. PCL-5 scores were recorded before treatment, 7 days after treatment, and at 30 day follow-up. Although PTSD symptoms significantly and robustly improved over time [before treatment, M=46.6,sd=13.35 end of treatment (7 days after infusion): M=33.8, sd=18.37; 30-day follow-up: M=30.79,sd=15.79] there was no significant difference between the ketamine and midazolam groups in the rate of improvement or in the PTSD score at the end of treatment and follow-up. Using Bayesian method (see Methods) revealed a significant difference between the scores before treatment to the other time points. In order to assess the statistical difference, we have compared the posterior distribution of each of the two time-points (i.e. 7 and 30 days after infusion) to the posterior distribution of scores before treatment. This revealed that the overlap of the other two distributions (after treatment, and 30 days), with the first distribution (after treatment), was less than 0.001 percent, suggesting a robust effect.

## Discussion

The goal of this study was to use a biomarker, the amygdala, as a main target engagement region, to test the ability of ketamine to enhance post-retrieval extinction of original traumatic memories associated with PTSD. We hypothesized that ketamine would reduce subsequent neural and emotional reactivity to the memories following systemic exposure. Further, we aimed to assess the extent to which ketamine effects were produced by interfering with reconsolidation or enhancement of extinction.

Our data provide evidence of a prominent ketamine effect, which was characterized by a pattern of alterations in circuit function that were more consistent with reconsolidation post-retrieval extinction pathway than with enhancement of classic extinction. We found that the ketamine group showed lower amygdala reactivity to recall of subjective traumatic events compared to the midazolam group, with no change in connectivity between the amygdala and vmPFC. Moreover, connectivity between the amygdala and posterior-hippocampus was reduced in the ketamine, but not the midazolam, group after treatment. Lastly, reduced physiological arousal, measured by skin conductance response, was found in the ketamine group but not in the midazolam group.

The amygdala is a key node in neural networks engaged in extinction and post-retrieval extinction processes ^1,6,8,11^. Some studies argue that diminished amygdala activation is associated with post-retrieval extinction, which is in line with the classic reconsolidation findings, blocking protein synthesis in the basolateral amygdala (BLA)^1^. Other studies have found that extinction learning involves an inhibitory signal from the prefrontal cortex, mainly the vmPFC to the amygdala and hippocampus^6^. Taken together, these findings suggest that extinction learning should present with higher connectivity between vmPFC and amygdala and higher activation of the vmPFC. Reconsolidation-based extinction (i.e. post-retrieval extinction), on the other hand, should present with lower activation in the amygdala, regardless of vmPFC-amygdala connectivity. Our findings of diminished activation of the amygdala and lack of higher connectivity between vmPFC and amygdala, in response to trauma recall in the ketamine group, are consistent with the post-retrieval extinction theory^10^.

NMDARs are required for the transition of memory from a fixed to a labile state. For example, an NMDAR antagonist, but not an AMPAR antagonist, blocked the reconsolidation process ^18^. NMDAR activation may be upstream of the stimulation of protein synthesis in the mechanistic cascade responsible for making memories labile, as the addition of anisomycin does not augment the interference with reconsolidation produced by an NMDAR antagonist ^1,18^. Midazolam, a benzodiazepine positive allosteric modulator of GABA_A_ receptors, has also been reported to interfere with the reconsolidation of fear ^45,46^. Our study, which employed doses of midazolam and ketamine selected for their similar tolerability^47^, found substantially greater post-retrieval extinction in participants who received ketamine than in those who received midazolam. This finding suggests that the transition of memory to a labile state is relatively more sensitive to disruption of NMDAR signaling than reductions in excitability produced by facilitating GABA_A_ receptor signaling. Although NMDAR antagonists acutely reduce neuroplasticity, ketamine, 24 hours after acute subanesthetic dose, promotes neuroplasticity by enhancing glutamate release, raising brain-derived neurotrophic factor (BDNF) levels, activating mTORC1 signaling, producing cortical synaptogenesis, and stimulating hippocampal neurogenesis ^19,20^, all of which are important in reconsolidation processes ^23,24^. Supporting this hypothesis, ketamine-related increase in hippocampal BDNF gene expression reduces reactivation of fear memories in rats^48^. The well-known antidepressant ^49,50^ and anxiolytic^51^ effects of ketamine also emerge only several hours to several days after ketamine administration.

Our results also show reduced amygdala-hippocampal connectivity following ketamine infusion. This effect was only apparent 7 days after infusion and not in the follow-up session (30 days), suggesting a transient effect. Some studies have demonstrated the relevance of dorsal amygdala-hippocampus connectivity in contextual fear memory ^11,14^, with some associating the level of amygdala - hippocampus connectivity to the long term retention of fear learning ^15^. In a recent study, Ressler and colleagues showed that the hippocampus was involved in contextual fear memory and that inhibition of protein synthesis in hippocampal ensemble disrupted the reconsolidation of fear memories^52^. The authors argue that these findings might help us better understand real-life traumatic memories (which we study here). The lower hippocampal reactivity to traumatic memory in the ketamine group, as well as the lower hippocampus-amygdala connectivity, might be a presentation of these findings in humans, on naturalistic traumatic memories. Several other studies found that the level of connectivity between the amygdala and hippocampus is associated with the ability to encode long-term memories ^53–57^. One study showed decreased amygdala insulo-temporal region resting-state connectivity in MDD patients, using magnetoencephalography (MEG;^58^. A recent study found that higher amygdala-hippocampus connectivity was positively associated with enhanced memory under stress, but not neutral, conditions, in healthy individuals^56^. Based on these findings, it is reasonable to assume that the decreased connectivity between the amygdala and posterior hippocampus found in our study is another biomarker of decreased fear memory trace to support the enhancement of memory reconsolidation using NMDA.

The facilitation of post-retrieval extinction of trauma memories in this study raises the possibility that behavioral interventions might enhance the reported efficacy of ketamine in the treatment of PTSD^59^. PTSD is a highly disabling disorder with few effective pharmacotherapy options^60^. It is speculated that PTSD derives from overgeneralization of fear responses and excessive or maladaptive reconsolidation of the traumatic memories. As such, reconsolidation-based treatments can be clinically relevant for treating such a disorder. Although some evidence-based treatments for PTSD are highly effective, they reach 50% remission at best^61^. The idea of using ketamine as a facilitator is consistent with a recent study conducted in alcohol use disorder patients^62^. In this study, ketamine was ineffective in reducing drinking by itself. However, when administered in conjunction with alcohol cues, ketamine interfered with the reconsolidation of alcohol-reward-related memories and significantly reduced drinking.

A self-report measure of PTSD symptom severity failed to separate the effect of ketamine and midazolam in symptom reduction. It is possible that a subjective account of psychological distress might be less sensitive to change than objective psychophysiological and neurobiological measures. This assumption is in line with the NIMH Research Domain Criteria (RDoC) initiative to further establish biomarkers associated with mental disorders.

This study has some limitations. First, the number of participants in each group is relatively modest, and larger studies will need to replicate the findings. This concern is partially mitigated by the longitudinal aspect of the study, as well as the findings, which are consistent with other animal and human studies. Another potential limitation is the use of an active psychotropic drug, midazolam. We used midazolam as a comparator to control for the subjective effects of ketamine and to preserve the blind conditions. This limits our inferences, however, to the relative effects of similarly tolerable doses of ketamine and midazolam. As midazolam may also interfere with reconsolidation, the prominent impact of ketamine versus midazolam in this study attests to the robustness of the ketamine effects. In order to fully unfold the effect of ketamine, a control group receiving ketamine without recall of trauma is needed.

Notwithstanding these limitations, the present study finds that one-time ketamine infusion might enhance post-retrieval extinction of traumatic memories. This is apparent in the effects found in the ketamine vs. the midazolam group: diminished amygdala reactivity to recall of traumatic events, attenuation of connectivity between amygdala and hippocampus, and hippocampus and vmPFC, and attenuation of skin conductance response to the recall of traumatic events. These findings replicate previous findings in both animal models and humans, on a wider spectrum of psychiatric disorders and both appetitive and aversive memories. As the enhancement of post-retrieval extinction presented here was demonstrated using real-life traumatic events, the applicability of this procedure is high and it might serve as a potential novel future intervention for PTSD and anxiety disorders.

## Data Availability

Data and analysis can be found in the github repository. All raw data are available from the corresponding author upon reasonable request.
Code availability:
All analysis scripts can be found here https://github.com/orduek/KPE.

https://github.com/orduek/KPE

## Acknowledgement

The main source of funding for this work was provided by: Independent Investigator Grant from the Brain and Behavior Research Foundation (IHR); by the Clinical Neurosciences Division of the National Center for PTSD (IHR);a donation from the American Brain Society (IHR), and the Yale Center for Clinical Investigation(YCCI) supported by CTSA Grant from the National Center for Advancing Translational Science (NCATS), a component of the National Institutes of Health (NIH). The contents are solely the responsibility of the authors and do not necessarily represent the official view of NIH

## Author Contributions

I.H.R designed the study. O.D., B.K., S.A., C.G., M.M and I.H.R. collected the data. O.D. and Y.L. analyzed the data, I.L and I.H.R. contributed to the data analyses. J.K. has contributed to the interpretation of the results. O.D., I.L. and I.H.R. wrote the first draft of the manuscript. All authors contributed to the final version of the manuscript.

## Data availability

Data and analysis can be found in the github repository. All raw data are available from the corresponding author upon reasonable request.

## Code availability

All analysis scripts can be found here https://github.com/orduek/KPE.

## Footnotes

1 HDI refers to High Density Interval. Instead of a confidence interval that assumes normal distribution, HDI will end with the relevant interval in any kind of distribution. For more information, please see https://www.sciencedirect.com/topics/mathematics/highest-density-interval

