## Supplement 1 for "Modulating amygdala activation to traumatic memories with a single ketamine infusion"

### Supplement 1: DiFuMo regions extracted

**Amygdala:**

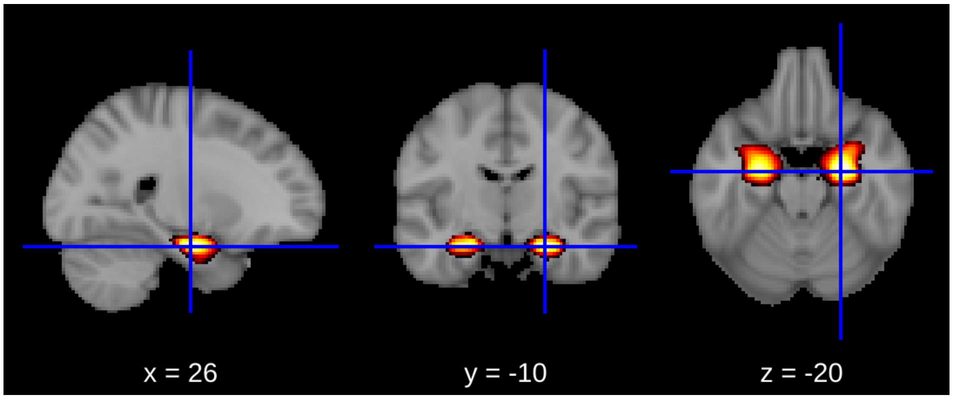


**Anterior hippocampus:**

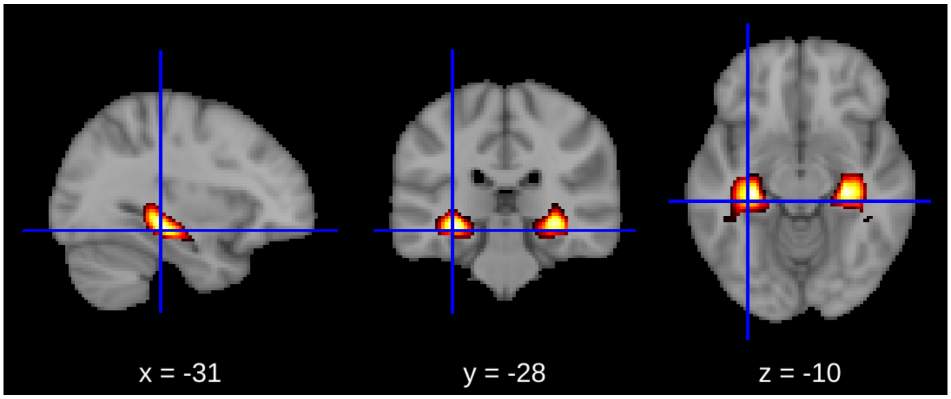


**Posterior hippocampus:**

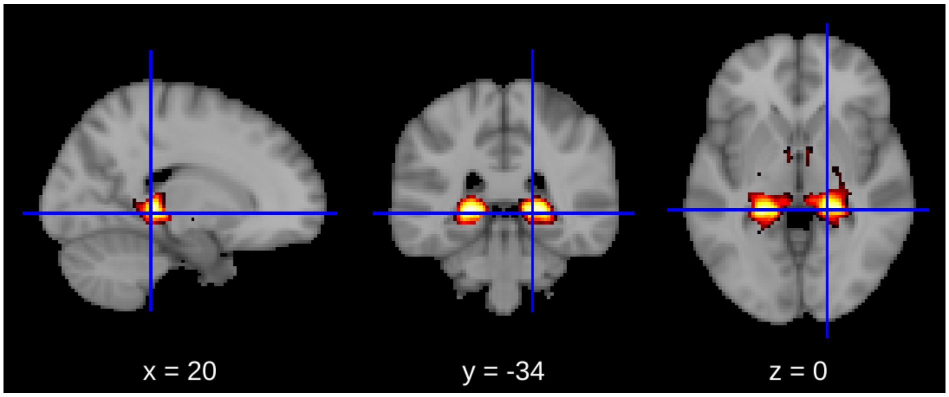

**Anterior vmPFC:**

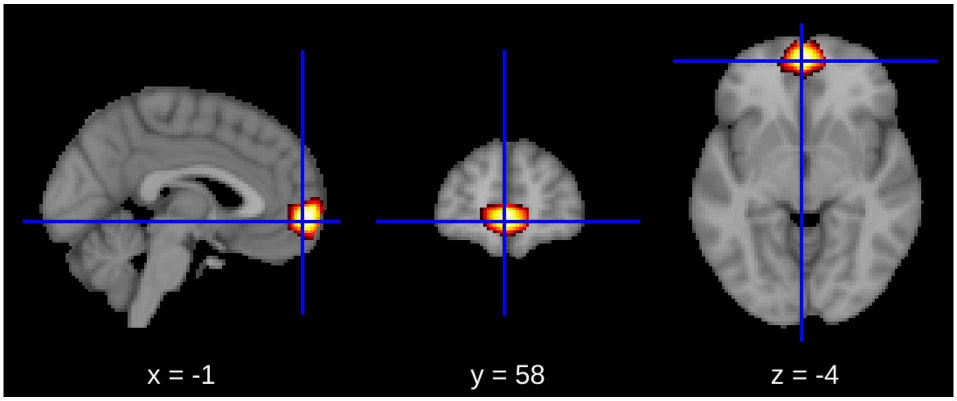


**vmPFC:**

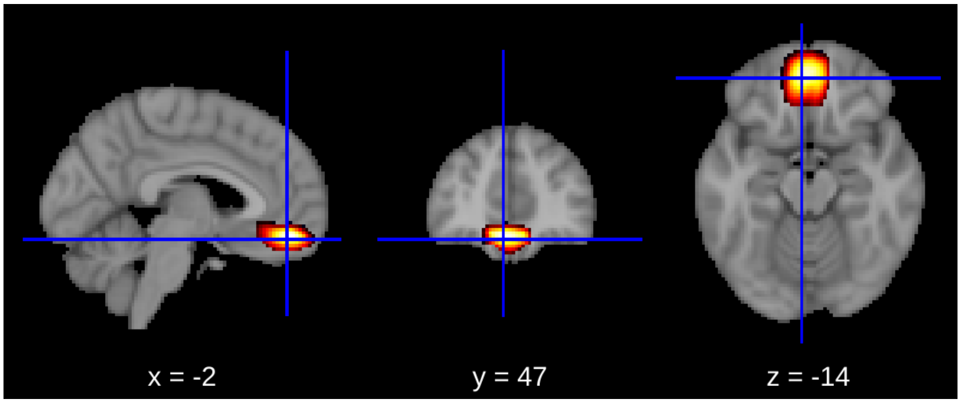
