## Supplement 2 for "Modulating amygdala activation to traumatic memories with a single ketamine infusion"

**vmPFC activation results:**

Before treatment, average vmPFC reactivity to trauma vs neutral script was -0.03 (sd=0.75) in the ketamine group [N=13] and -0.03 (sd=0.54) in the midazolam group [N=13]. At the end of treatment (7 days after infusion) the average vmPFC reactivity was -0.03 (sd=0.39) in the ketamine group [N=13], and 0.07 (sd=0.46) in the midazolam group [N=12]. At 30-days follow-up the vmPFC reactivity was -0.01 (sd=0.48) in the ketamine group [N=12] and 0.01 (sd=0.34) in the midazolam group [N=10]. The statistical analysis showed no significant effect for time, also comparing between ketamine and midazolam groups (in each time point), revealed no difference. Figure 5 presents the average vmPFC reactivation in each group across the different time points.

#### Symptoms change

**Using Mixed models:** A linear mixed model, using subjects nested in experimental group as random effect, indicated only a significant fixed effect for time ($F[2,40]=19.3$, $p<0.0001$) with no effect for medication ($F[1,20]=0.39$, $p=0.54$) or medication x time interaction ($F=1.3$, $p=0.28$).

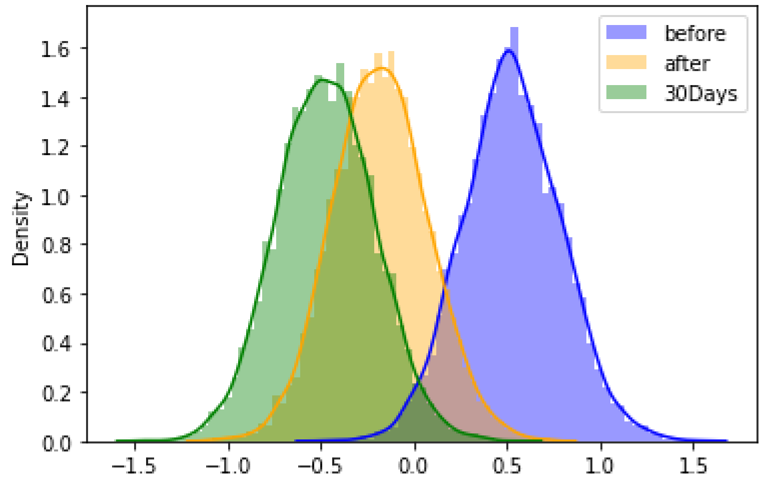


The figure presents the posterior distributions of the slope from the Bayesian model of PCL-5 scores. Before is the coefficient before treatment; after = 7 days after infusion; 30Days = 30 days after infusion.

### Sad vs. Neutral contrast results:

#### Changes in neural activation

**Amygdala**:

Comparing the amygdala activation while listening to the sad script (vs. neutral) revealed no groups differences before [ketamine M=0.19 (sd=0.26], midaolzam: 0.11 (sd=0.36), mean posterior=0.08 (sd=0.13) 90%HDI(-0.136,0.299)]; after [ketamine M=0.11 (sd=0.29], midaolzam: 0.08 (sd=0.35), mean posterior=0.028 (sd=0.14) 90%HDI (-0.196,0.268)]; and in 30-day follow-up [ketamine M=0.01 (sd=0.24], midaolzam: -0.04 (sd=0.28), mean posterior = 0.059 (sd=0.12) 90%HDI (-0.158,0.254)]. Although no difference was found between the groups, there was an effect for time. Comparing the amygdala activation of both goups in the 30-day followup to the one before treatment revealed a significant decline in activation (overlapping of posterior distiburions was 0.04). There was no significant decline 7-days after infusion (see figure below).

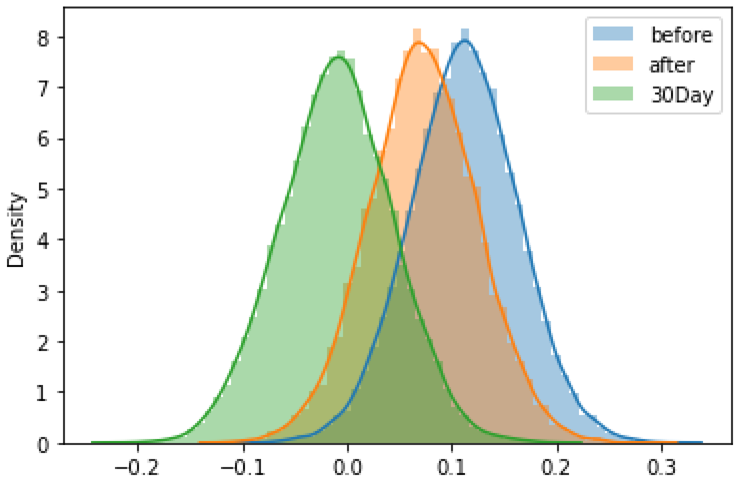


The figure portrays the posterior distributions of the time coefficients (of amygdala activation [sad> neutral] before treatment, after treatment, and in 30-day follow-up.

**Hippocampus:**

Comparing the hippocampus activation while listening to the sad script (vs. neutral) revealed no groups differences before [ketamine M=0.15 (sd=0.34], midaolzam: 0.21 (sd=0.40), mean posterior=-0.06 (sd=0.15) 90%HDI(-0.325,0.187)]; after [ketamine M=0.12 (sd=0.27], midaolzam: 0.01 (sd=0.37), mean posterior=0.108 (sd=0.14) 90%HDI (-0.139,0.327)]; and in 30-day follow-up [ketamine M=-0.07 (sd=0.32], midaolzam: -0.05 (sd=0.24), mean posterior = -0.01 (sd=0.13) 90%HDI (-0.239,0.208)]. Although no difference was found between the groups, there was an effect for time. Comparing the hippocampus activation of both goups in the 30-day followup to the one before treatment revealed a significant decline in activation (overlapping of posterior distiburions was 0.01). There was no significant decline 7-days after infusion (see figure below).

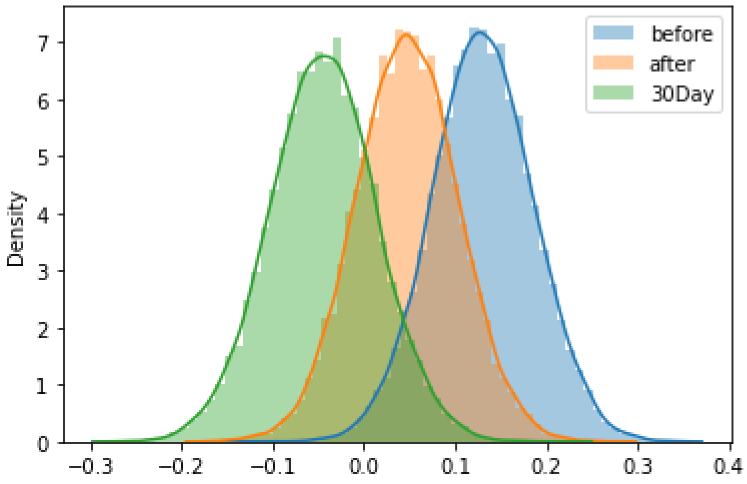


The figure portrays the posterior distributions of the time coefficients (of hippocampus activation [sad>neutral] before treatment, after treatment, and in 30-day follow-up.

**vmPFC:**

Comparing the vmPFC activation while listening to the sad script (vs. neutral) revealed no groups differences before [ketamine M=0.24 (sd=0.46], midaolzam: 0.24 (sd=0.27), mean posterior=-0.003 (sd=0.16) 90%HDI(-0.244,0.264)]; after [ketamine M=0.06 (sd=0.53], midaolzam: 0.09 (sd=0.51), mean posterior=-0.026 (sd=0.22) 90%HDI (-0.410,0.330)]; and in 30-day follow-up [ketamine M=-0.04 (sd=0.45], midaolzam: 0.04 (sd=0.31), mean posterior =

-0.086 (sd=0.19) 90%HDI (-0.377,0.237)]. No difference was found between timepoints in the activation of hippocampus as well. see figure below.

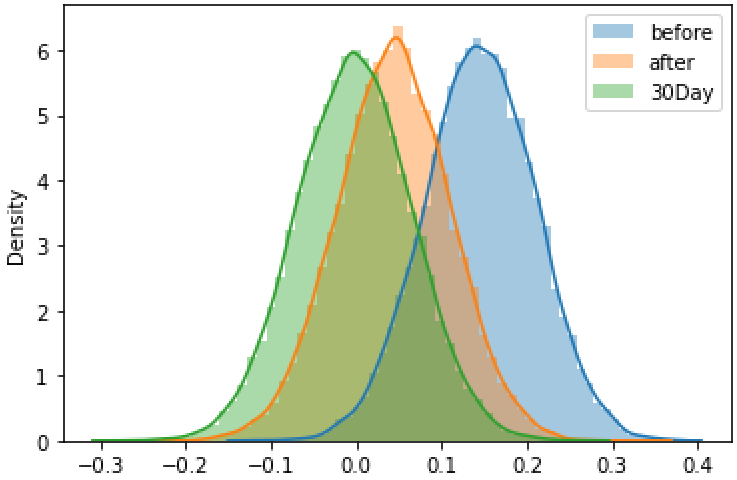


The figure portrays the posterior distributions of the time coefficients (of vmPFC activation [sad>neutral] before treatment, after treatment, and in 30-day follow-up.

**<u>Changes in connectivity:</u>**

**Amygdala-vmPFC:**

amygdala-vmPFC functional connectivity was not significantly different between the groups before [ketamine M=0.39 (sd=0.29], midaolzam: 0.52 (sd=0.18), mean posterior=-0.135 (sd=0.11) 90%HDI(-0.308,0.04)]; after [ketamine M=0.44 (sd=0.16], midaolzam: 0.47 (sd=0.16), mean posterior=-0.024 (sd=0.07) 90%HDI(-0.146,0.096)]; and in 30-day follow-up [ketamine M=0.37 (sd=0.24], midaolzam: 0.49 (sd=0.19), mean posterior=-0.123 (sd=0.10) 90%HDI(-0.292,0.048)]. There was no time effect as well.

**Amygdala - posterior hippocampus:**

Amygdala-posterior hippocampus functional connectivity was not significantly different between the groups before [ketamine M=0.25 (sd=0.26], midaolzam: 0.23 (sd=0.31), mean posterior=0.01 (sd=0.12) 90%HDI(-0.195,0.200)]; after [ketamine M=0.22 (sd=0.24], midaolzam:

0.28 (sd=0.29), mean posterior=-0.054 (sd=0.11) 90%HDI(-0.241,0.132)]; and in 30-day follow-up [ketamine M=0.28 (sd=0.23], midaolzam: 0.24 (sd=0.30), mean posterior=0.049 (sd=0.12) 90%HDI(-0.129,0.271)]. There was no time effect as well.
